# Digital Health for Vulnerable and Disabled Populations in Natural Disaster and Conflict Settings: A Scoping Review Contributing to Sustainable Development Goals 3, 10, 11 and 13

**DOI:** 10.1101/2025.09.15.25335819

**Authors:** Amna Zia, Anam Zafar, Saima Riaz, Muhammad Naveed Babur, Samrood Akram

**Affiliations:** Department of Physical Therapy and Rehabilitation Sciences, Superior university, Lahore; Department of Physiotherapy, Mayo Hospital, Lahore, Pakistan; Department of Physical Therapy and Rehabilitation Sciences, Superior university, Lahore, Pakistan; Department of Physical Therapy and Rehabilitation Sciences Faculty of Allied Health Sciences Superior university, Lahore, Pakistan

**Author notes:** Conceptualised the study, led the writing, contributed to screening, data charting, and thematic synthesis, coordinated the scoping process (Amna Zia). Contributed to screening and data charting (Anam Zafar). Supervised the overall review, provided methodological oversight, contributed to manuscript review and editing (Saima Riaz). Supervised the overall review, ensured critical revisions and academic integrity (Muhammad Naveed Babur). Contributed to manuscript editing (Samrood Akram).

**Keywords:** Artificial intelligence, Conflict, Digital health, Disability, Natural Disaster, Telemedicine, Telerehabilitation, War

## Abstract

**Background:** People affected by conflict, war, and climate-related disasters, particularly those living with disabilities, often face the most significant barriers to accessing essential health services. Despite their heightened vulnerability, healthcare in these settings is frequently fragmented and difficult to access. In recent years, digital health interventions have been increasingly explored as practical tools to bridge these gaps and enhance service delivery in crisis contexts. However, there remains limited clarity on how effectively these interventions serve vulnerable and disabled populations. This scoping review aimed to examine the nature, scope, and reported outcomes of digital health interventions implemented in natural disaster and conflict settings, and to assess how these efforts align with Sustainable Development Goals (SDGs) 3, 10, 11, and 13.

**Methods:** A comprehensive search was conducted across PubMed, EBSCOhost, and the Cochrane Library for studies published up to May 2025. The search strategy targeted digital health interventions involving vulnerable populations in the context of disasters or armed conflict. Following the PRISMA-ScR guidelines screening process was done systematically. Guided by the Arksey and O’Malley scoping review framework, relevant data were extracted that captured study characteristics, population details, type of digital intervention, reported outcomes, and contextual barriers or enablers. The extracted data were then analysed thematically to identify key patterns across studies.

**Results:** Eight studies met the inclusion criteria, encompassing a range of designs including narrative reviews, qualitative case studies, a clinical trial, and a scoping review. The interventions covered telemedicine, telerehabilitation, mobile health platforms, virtual mental health therapies, and remote problem-solving treatments. While many studies reported improved healthcare access, continuity of care, or system resilience, few offered detailed outcomes specific to persons with disabilities. Common enablers included public-private partnerships, trained staff, and scalable technologies. Barriers involved infrastructure damage, digital literacy gaps, and weak governance. Most interventions were reactive, with limited evidence of long-term integration or inclusive design.

**Conclusions:** Digital health interventions have potential to support vulnerable and disabled populations during crises, contributing to SDGs related to health, equity, urban resilience, and climate action. However, inclusion remains uneven, and most studies lack detailed evaluation and long-term sustainability planning.

## Introduction

Over the past few decades, the rate and magnitude of natural disasters and armed conflicts have been aggravated by occurrence of climate change, high urbanisation, and weak socio-political conditions (1, 2, 3, 4). The occurrence of such events destabilizes the health system, destroys infrastructure, and overloads humanitarian services (5, 6, 7). Most importantly, they affect the vulnerable groups disproportionately as pointed to the broad definition of vulnerable called in this review people whose health, mobility or access to care has been impaired due to environmental, structural or social factors. This includes, but is not limited to, persons with disabilities (PWD), displaced populations, individuals with chronic conditions, older adults, military personnel affected by conflict, and those living in remote or under-resourced settings.

Globally, over 3.6 billion people live in areas highly vulnerable to climate change, and climate-related health risks could cause an additional 250,000 deaths annually between 2030 and 2050 due to undernutrition, malaria, and heat stress (8). These challenges are all exacerbated in conflict regions, particularly due to poor governance and damaged infrastructure, which compound difficulties in care delivery (1, 5). During these situations, there can be a considerable lack of access to healthcare- especially among the populations who use regular care, rehabilitation, or assistive technology (9, 10).

Such digital health tools as telemedicine, mobile health (mHealth) apps, electronic health records (EHRs), remote monitoring, and AI assisted systems have become scalable solutions in times of crisis (11). These technologies have the promise of supporting continuity of care, building resiliency, and accessing the underserved population who would not receive traditional health services (12). They have had encouraging use in humanitarian contexts but even with these innovations, there still exist gaps. Many interventions are not designed with inclusivity in mind. The situation can be described by the fact that over 1.3 billion people worldwide live with a disability, of which 80 percent live in low- and middle-income countries , only 15 percent of countries incorporate disaster risk reduction (DRR) strategies to integrate people with disabilities (13, 14). Furthermore, 86% of persons with disabilities report exclusion from community-level disaster preparedness planning (15) and similar patterns of neglect affect other vulnerable groups as well (16).

Digital health interventions, therefore, offer a critical opportunity to close these equity gaps. They are emerging as powerful tools to enhance healthcare access for marginalised populations, particularly in disaster- and conflict-affected settings where conventional health systems are often disrupted (17). Despite commitments to equity and inclusion under global frameworks like the Sustainable Development Goals (SDGs), there remains limited consolidated evidence on how digital health technologies are being utilised to support vulnerable populations during emergencies. This scoping review aims to map the scope, nature, and characteristics of existing research in this area and to examine the extent to which digital health interventions contribute to SDGs 3 (good health and well-being), 10 (reduced inequalities), 11 (sustainable cities and communities), and 13 (climate action). To date, no comprehensive review has explored this crucial intersection.

## Methods

### Title and Protocol Registration

The protocol for this scoping review was registered in the Open Science Framework (OSF) https://osf.io/y2ta3 (18).

### Study Design

The study was based on the six-stage scoping review methodological framework provided by Arksey and OMalley (2005) and refined by Levac et al. (2010) (19). The reporting is done according to PRISMA Extension for Scoping Reviews (PRISMA-ScR) (20). The objective of this review was to systematically investigating the use of digital health interventions in support of vulnerable and disabled populations during disasters and conflict, and also evaluating how digital health interventions can support Sustainable Development Goals (SDGs) 3, 10, 11 and 13.

### Stage 1: Identifying the Research Question

The primary research question for this review was:

**What types of digital health interventions have been used to support the health, resilience, and inclusion of vulnerable and disabled populations during natural disasters and conflicts and how do these interventions contribute to the achievement of SDGs 3, 10, 11, and 13?**

This question was intentionally framed to cover a wide range of digital solutions while maintaining a clear link to global health and development priorities. The aim was to explore not only the character of digital interventions in such high-risk environments but also their performance and extended implication around equity, resilience, and sustainable distributive wellbeing.

To frame the research question, the Population–Concept–Context (PCC) framework was applied. The specified population included vulnerable groups such as individuals affected by conflict or natural disasters, persons with disabilities, and military personnel. The central idea was the digital health intervention as telemedicine, mHealth, telerehabilitation services, and digital mental health services. The setting was focused on natural hazards (floods, earthquakes, wildfires), armed conflicts or wars that are identified to interfere with the conventional healthcare system.

### Step 2: Identifying Relevant Studies

An extensive search strategy was designed and used by searching three databases including PubMed, Cochrane Library, and EBSCOhost. All databases were searched between April and June, 2025. The PubMed search (shown in **Table 1**) served as a base and was adapted to suit each database. Detailed strategies for other databases are attached in supplementary fie. The search timeframe spanned from 2000 to 2025 to capture the rise of digital health innovation in disaster and emergency response.

**Table 1:** Search strategy used for PubMed.

| Search # | Query Summary | Results |
| --- | --- | --- |
| #1 | (((((climate) OR (weather)) OR (Storme)) OR (fire)) OR (earthquake)) OR (natural disaster)) OR (flood)) OR (wildfire)) OR (tornado) | 1,011,162 |
| #2 | (((((disability) OR (mental disability)) OR (physical disability)) OR (person with disability)) OR (handicap)) OR (disabled)) OR ("differently abled")) OR (war) | 561,970 |
| #3 | (((((telehealth) OR (telemedicine)) OR (Artificial intelligence)) OR (digital health)) OR (eHealth)) OR (mHealth)) OR (assistive technologi*) OR (remote monitoring) | 527,958 |
| #4 | (((((((((climate) OR (weather)) OR (Storme)) OR (fire)) OR (earthquake)) OR (natural disaster)) OR (flood)) OR (wildfire)) OR (tornado)) AND ((((((disability) OR (mental disability)) OR (physical disability)) OR (person with disability)) OR (handicap)) OR (disabled)) OR ("differently abled")) OR (war)) AND ((((((telehealth) OR (telemedicine)) OR (Artificial intelligence)) OR (digital | 174 |
|  | health)) OR (eHealth)) OR (mHealth)) OR (assistive<br>technologi*) OR (remote monitoring)) |  |

To ensure a comprehensive search, we combined keywords reflecting the main concepts of digital health, telemedicine, mHealth, disability, vulnerable populations, conflict, and disaster. We used Boolean operators like AND and OR to structure the search logic, and applied truncation to include different word forms. Where available, MeSH terms were also incorporated to improve the accuracy and depth of the search across databases.

Google Scholar, reports from WHO, UNDRR, UNICEF, NGO websites (e.g., Handicap International, CBM, Sehat Kahani), and manual backward citation tracking were also used to identify additional relevant literature that may not have been captured through database searches.

This method was framed to enable maximum coverage in terms of disciplines covering subjects like public health, rehabilitation, technology, and humanitarian studies and at the same time keep relevance to the main research question.

### Stage 3: Study Selection

The study selection process is shown in the PRISMA flowchart (**Figure 1**). The screening was done after eliminating the duplicated and irrelevant records in two steps. Initially, titles and abstracts were examined in terms of relevance. Articles whose reports did not specifically focus on use or effects of digital health in disasters or conflicts settings, were excluded. Full texts of potentially relevant articles were then evaluated using pre-determined inclusion and exclusion criteria in the second step.

### Inclusion Criteria

- Digital health interventions (e.g., telemedicine, mHealth, eHealth, telerehabilitation)
- In disaster or conflict settings
- Focused on persons with disabilities (PWD), vulnerable populations (Military persons (active or veteran), People in the war zone/Conflict setting)
- Primary studies, Published Reviews (systematic, scoping or narrative), or case studies/reports
- English language
- Published between 2000–2025

### Exclusion Criteria

- No digital health component
- No mention of disaster/conflict setting
- Opinion pieces, editorials, or short commentaries

After this procedure, eight studies were fulfilled the inclusion criteria to be subjected to data extraction and analysis. These were narrative, a scoping review, case studies and a randomised controlled trial.

### Stage 4: Charting the Data

An extraction table with a structure of data was designed and iteratively improved. In eight studies, the following variables were extracted: Name(s) of author(s) and year of publication, study design, study objective, target population, type of digital health intervention, outcome, enables/barriers and relevant SDG contribution (3, 10, 11, and 13).

This procedure offered a systematic means of recording not just the technologies used but also its success, and the situational barriers and promoters that affected the final result.

### Stage 5: Collating, Summarising and Reporting the Results

Thematic synthesis was used to analyse the findings across studies. Given the diversity in methods and settings, a narrative approach was chosen to allow for the identification of cross-cutting themes without losing contextual detail. Three key domains emerged: types and functions of digital health interventions, barriers and enablers to implementation and impact on equity, resilience, and SDG alignment.

### Stage 6: Stakeholder’s consultation

Although such involvement of stakeholders in scoping review is a worthy feature, it was not implemented because of time and financial limitations. That is why the results are grounded only on published academic and grey literature.

However, the importance of engaging with end-users, especially persons with disabilities, frontline healthcare workers, and humanitarian agencies, is acknowledged.

## Results

A total of 604 records were initially identified through database searches: PubMed (n = 174), EBSCOhost (n = 422) and the Cochrane Library (n = 8). Following the removal of 502 unrelated or duplicated records, 102 records were screened based on title and abstract. Out of which, 58 were removed as they were irrelevant or unavailable as full text.

The other 44 full-text reports were evaluated for eligibility. Following a detailed review, 36 reports were excluded for one or more of the following reasons: the study did not focus on disaster or conflict settings, lacked relevance to vulnerable or disabled populations, or was an editorial, commentary, or short communication. Ultimately, eight studies met all inclusion criteria and were included in the final synthesis. The selection process is summarised in the PRISMA flow diagram (**Figure I**).

**Figure 1:**
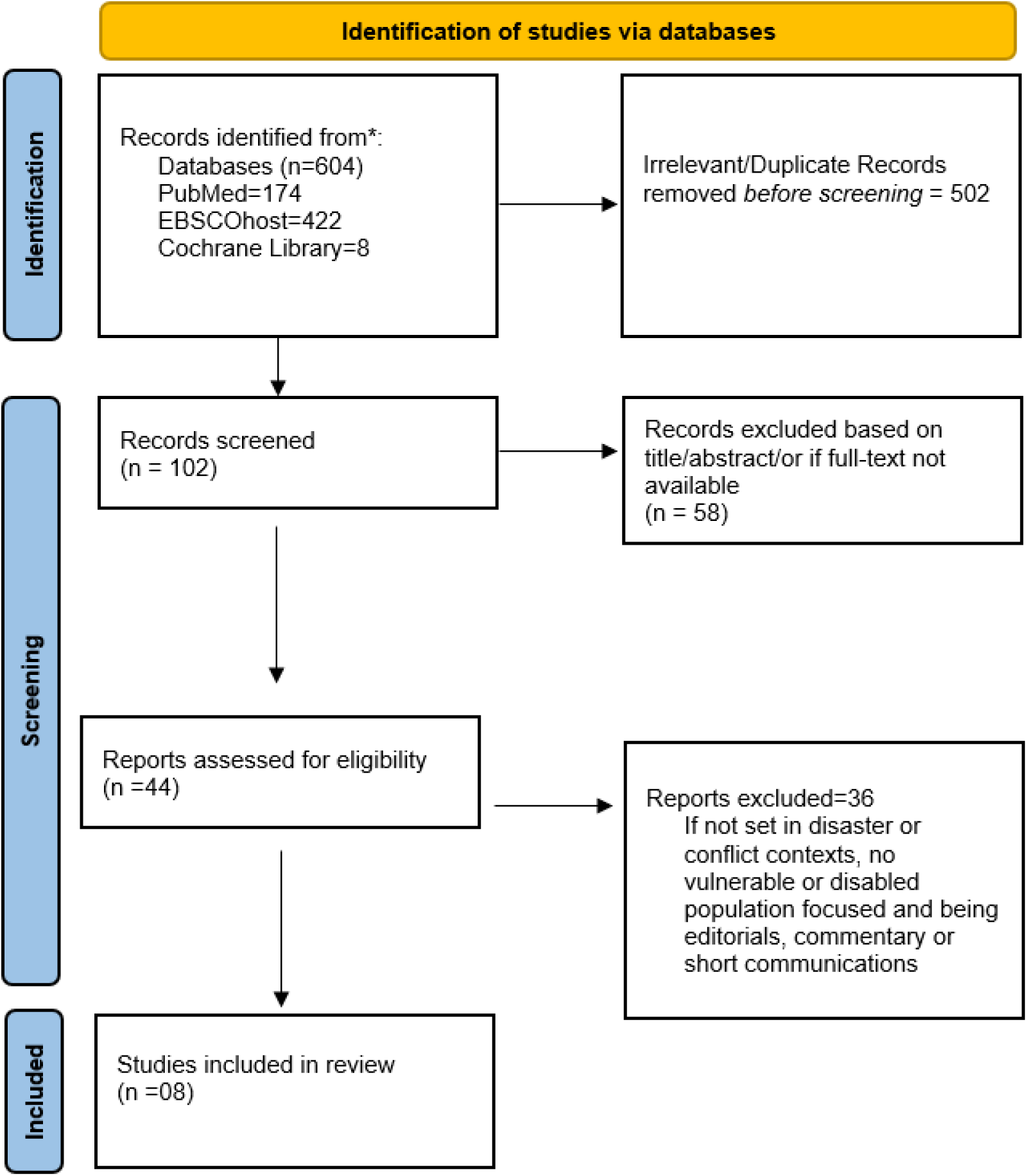
PRISMA Flowchart.

### Included Studies Characteristics

The eight included studies in this review covered a wide variety of methodological grounds, contexts, and demographic groups, with all that evidence the variety of digital health interventions utilized in the setting of natural disasters and conflicts. Summary of the included studies is shown in **Table II**. The types of studies included three narrative review articles (21, 22, 23) , one scoping review (24), one descriptive case study (25), one qualitative case study (26), one mixed-methods review (27), and one randomised clinical trial (28). This review diversity in study designs allowed the wide synthesis of empirical and theoretical findings on the topic of the digital health response to humanitarian situations.

**Table II:** Digital Health Interventions for Vulnerable and Disabled Populations in Disaster and Conflict Settings.

| Author (Year) | Study Objective | Study Design | Population | Digital Intervention | Outcomes | Barriers/Enablers | Conclusion |
| --- | --- | --- | --- | --- | --- | --- | --- |
| **Ugwu et al. (2025) (23)** | To explore how SDG-aligned digital health strategies improve healthcare in conflict zones | Narrative review | Conflict-affected populations in LMICs | Telemedicine, mHealth apps, mobile clinics, EHRs, CHWs | Improved access and continuity of care; integrated community-based and mental health interventions | Enablers: Technology integration, community engagement, multi-sectoral partnerships<br>Barriers: Governance instability, limited infrastructure, fragmented systems | Digital health tools, when aligned with SDGs 3 offer a strategic pathway to strengthen health system resilience and access to care in fragile, conflict-affected settings, especially among vulnerable populations. |
| **Khuld (2024) (26)** | To assess Sehat Kahani's telemedicine model during floods in Pakistan and its impact on health outcomes and system resilience | Qualitative case study | Flood-affected communities in Sindh and Balochistan; healthcare providers, funders, management | Telemedicine mobile units, physical camps, telehealth app ("One Window") | 25,000+ patients reached (including women, children, elderly); timely diagnosis and treatment; reduced mortality | Enablers: Public-private partnerships, scalable tech, trained staff, HIPAA-compliant systems<br>Barriers: Connectivity limitations, patient privacy, infrastructure strain | Multi-tier telehealth improved healthcare access and disaster response; model is replicable in disaster-prone, resource-limited settings. It supports SDGs 3, 10, 11, and 13 by addressing health disparities, strengthening emergency response, and promoting adaptive digital health strategies in vulnerable communities. |
| **Lokmic-Tomkins et al. (2024) (27)** | Map lessons from natural disasters around digital health | Mixed-methods Review | Crisis-affected populations | Digital platforms (telemedicine, EMRs, citizen science) | Lessons on preparedness, resilience, health system adaptation | Barriers: lack of coordination, infrastructure gaps | Digital health supports climate-resilient and adaptive care models in disasters, with lessons informing future preparedness aligned with SDGs 3, 11, and 13; however, implementation depends on overcoming infrastructural and coordination challenges. |
| **Emük et al. (2023) (22)** | To explore the role and feasibility of telerehabilitation in post-earthquake settings | Narrative Review | Earthquake-affected individuals and communities | Telerehabilitation (video, mobile, online PT) | Enhanced access to care, reduced costs, remote management of physical and psychological impairments, support for triage and recovery | Enablers: Remote accessibility, cost-effectiveness, potential for psychological support<br>Barriers: Infrastructure damage, power/internet loss, lack of training, data privacy concerns | Telerehabilitation is a viable post-disaster solution to deliver timely and accessible rehabilitation care, but requires proper planning, infrastructure, and training for effective implementation.<br>It supports SDGs 3, 10, 11, and 13 by promoting health equity, resilience, and accessible care despite infrastructural challenges. |
| **Jones et al. (2020) (24)** | Review digital trauma therapies for PTSD | Scoping Review | Military, veterans, PSP with PTSD | Virtual CBT, CPT, PE via video platforms | Mixed evidence; stigma reduced; similar efficacy | Barriers: access, stigma; Enablers: remote reach | Virtual trauma therapies can improve PTSD care accessibility and reduce stigma among vulnerable populations; with context-sensitive design, they align with SDGs 3 by enhancing mental health service delivery. |
| **Pasipano dya & Shem (2020) (25)** | Report teleSCI use post-wildfire | Case Study | SCI adult in California wildfire | TeleSCI (SCI-focused telerehabilitation) | Maintained care, replaced equipment, emotional support | Barrier: wildfire loss; Enabler: pre-existing teleSCI program | TeleSCI enabled continuity of specialised rehabilitation during wildfires, illustrating how pre-established digital care systems can support SDG 3 by maintaining essential services for disabled individuals in emergencies. |
| **Bell et al. (2017) (28)** | To evaluate the efficacy of telephone-delivered problem-solving treatment (PST) in reducing psychological and physical symptoms among military personnel with combat-related mTBI | Randomised Clinical Trial | 356 post-deployment active-duty U.S. service members with confirmed mild TBI from Iraq/Afghanistan | Telephone-based PST (up to 12 sessions over 6 months) vs. Education-only (EO) group with brochures | PST significantly reduced psychological distress, depression, PTSD, and sleep disturbances at 6 months, but not post-concussive symptoms; no sustained effect at 12 months | Enablers: Participant preference for PST, remote access, reduced stigma; Barriers: dropout rate, short-lived impact, limited symptom severity in some | Telephone-based psychological support offered short-term mental health benefits for service members post-deployment, aligning with SDGs 3 by reducing psychological distress for individuals in conflict settings. |
| **Mesmar et al. (2016) (21)** | To explore how digital technologies were used to support health systems in conflict-affected settings in the Middle East and North Africa (MENA) region | Descriptive Review | Populations in conflict-affected MENA countries (e.g. Syria, Iraq, Palestine) | Digital health applications including eHealth and mHealth for surveillance, communication, and service delivery | Technologies improved disease surveillance, facilitated health service delivery, and supported provider coordination in insecure settings | Barriers: Infrastructure damage, lack of digital literacy, connectivity issues; Enablers: Growing mobile penetration, NGO involvement, urgency during conflict | Digital technologies enhanced service coordination and disease surveillance in conflict-affected MENA regions, contributing to SDGs 3, 10, 11, and 13 by supporting health system functionality amidst fragile, climate-stressed settings. |

Target populations varied considerably across the studies. Vulnerable groups included people living in conflict-affected areas of low- and middle-income countries (23), flood-displaced communities in Pakistan (26), earthquake survivors requiring rehabilitation services (22), and individuals with spinal cord injuries affected by wildfires (25). Other studies focused on psychological trauma among veterans, active-duty military personnel, and public safety personnel (24, 28). Mesmar et al. (2016) concentrated on populations across the Middle East and North Africa (MENA) region (21), while Lokmic-Tomkins et al. (2024) captured insights from multiple natural disaster settings globally (27). This wide range of affected populations underscores the adaptive potential of digital health in diverse sociopolitical and environmental conditions.

The types of digital health interventions that were reported in the literature were also diverse. They were telemedicine platforms (23, 26), the mobile health (mHealth) applications (28), electronic health records (23), telerehabilitation services (22, 25) and video or telephone-based virtual psychological therapies (24, 28), During flooding in Pakistan, Khuld (2024) tested a two tier telehealth system that included mobile medical units, physical field camps and a centralised telehealth application that provided care to more than 25,000 people (26). Likewise, Pasipanodya and Shem (2020) reported on how a teleSCI system adapted to uninterrupted specialist rehabilitation care of a person with an acquired spinal cord injury during a wildfire, offering direct rehabilitation care to an individual despite the overall destruction (25).

The results of these studies have focused on increased care access, care continuity, diminished psychological distress and enhanced health system resilience. In post-conflict scenarios, Ugwu et al. (2025) observed how digital health devices were in line with SDG 3 (Good Health and Well-being), where it has helped in providing integrated community-based and mental care, as well as technology integration and multi-sectoral collaboration to prop up institutional resilience (23). Khuld (2024) emphasized that scalable technologies and public-private partnerships played a critical role in providing the timely diagnosis and alleviating the mortality rates of the disaster-stricken regions (26). Bell et al. (2017) discovered that telephone-based problem-solving treatment decreased depressive symptoms, PTSD, and sleep impairments in military people with mild traumatic brain injury six months after treatment, but its positive effects were not found at one-year follow-up (28). In the meantime, Jones et al. (2020) stated that virtual trauma therapies, including cognitive processing therapy and prolonged exposure therapy, compared favorably to in-person versions and decreased stigmas related to mental health care although results were inconsistent and dependent on context (24).

The general barriers to implementation identified in the studies included infrastructural damage (21, 22), lack of internet and power connection (26), digital illiteracy, and data privacy issues (24). Indicatively, Emuk et al. (2023) observed that although telerehabilitation was effective in meeting both physical and psychological needs of post-earthquake recovery, scalability was threatened by technical and training deficiencies (22). On the contrary, successful implementation was enabled by a high engagement of communities (23), the presence of digital systems HIPAA-compliant (26), and digital infrastructures already established, which is the case in the wildfire case study Pasipanodya and Shem (2020) (25). Mobilisation of NGOs, cooperation with governmental agencies, and user-centred design was also indicated as critical success factors thereof in a number of interventions (21, 27).

Each of the included studies showed relevance with one or more of the Sustainable Development Goal (SDGs), but SDG3 (Good Health and Well-being) was the most widely discussed. The number of studies that fit in SDG 10 (Reduced Inequalities), embracing inclusive access to care in marginalised or disabled populations (26), SDG 11 (Sustainable Cities and Communities), empowering healthcare infrastructure and emergency response (27), and SDG 13 (Climate Action), and promoting climate-resilient care models (22). So, the body of literature demonstrates how digital health interventions can go beyond their direct service provision to influence the development of more equitable and sustainable health systems in fragile and disaster-prone settings.

### Thematic Analysis

This scoping review identified four interlinked themes reflecting the utility, challenges, and enablers of digital health interventions in disaster and conflict settings: access and continuity of care, health system resilience, equity and inclusion, and implementation barriers and facilitators.

### 1. Access to Care and Service Continuity

The access to healthcare through digital health greatly increased the availability of the healthcare system in areas where traditional systems were disrupted by disasters or conflicts. Telemedicine and mobile health platform served to maintain service provision among displaced and isolated populations.

Tthe telemedicine system of Sehat Kahani (mobile units, physical camps, and a centralised app) facilitated the treatment of more than 25,000 people, including women and elderly individuals, during the devastating floods in Pakistan (26). Ugwu et al. (2025) also showed the benefit of restoring the uncompleted care service by using telemedicine and EHRs and mHealth apps in post-conflict conditions, specifically in the case of maternal and mental health services, which comply with SDGs 3 and 16 (23).

Emulk et al. (2023) observed that telehealth services (telerehabilitation platforms) and access to mental health support in regions affected by earthquakes can eased the situations of in-person treatment, where it was impossible (22). Similarly, the paper by Pasipanodya and Shem (2020) outlined how a spinal cord injury patient in California was undergoing virtual consultations, equipment, and psychosocial care even after the California wildfires, due to a pre-existing teleSCI programme (25).

It was also reflected in studies specializing in the military. Bell et al. (2017) found that telephone-based therapeutic sessions showed positive impact on psychological improvements among traumatic brain injury patients in the military with mild injuries (28). A study by Jones et al. (2020) applied on rural veterans and safety staff found therapeutic processes of trauma treatment (e.g., CBT, PE) to be at least as effective as in-person therapies, and more accessible provider-wise and generally less judgmental (24).

Together, these results demonstrate the geographical, logistical, and social barriers mitigated by digital health, guaranteeing the continuity of care to vulnerable populations and are the direct contributions to SDG 3 and SDG 10.

### 2. Health System Resilience and Preparedness

Several studies highlighted how digital health contributes to systemic resilience, the ability to maintain essential functions during crises and adapt post-emergency. Incorporation of digital technologies in health governance enabled improved resource utilisation and diffuse responses.

By demonstrating the benefits of SDG-compatible digital tools to improve health provision across conflict areas by offering decentralised services and rapid exchange of information, even in conditions of governance breakdown, Ugwu et al. (2025) moderated the growing popularity of the notion of conflict zones as a source of emerging technologies (23). In a similar way, Lokmic-Tomkins et al. (2024) concluded that such tools as telehealth and EMRs assisted with the disaster preparedness and continuity of care in case of climate events, such as cyclones (27). However, they warned that it would require infrastructure, coordination and human capacity to implement.

Pasipanodya and Shem case studied showed the worth of well implemented systems, as it provided continuous care even during wildfires (25). Khuld (2024) documented that the HIPAA certification of Sehat Kahani, staff training, and public-private collaborations increased the sustainability of the Pakistani health system (26).

Mesmar et al. (2016) explained that in the MENA region eHealth and mHealth sustained communication and disease monitoring during conflict situations and was facilitated by NGO innovative solutions and mobile coverage, despite infrastructure and literacy issues (21).

These results are not just mobilizing the idea that digital health is a short-term solution but that it is also the foundation of stronger health systems ready to adapt to the future. This has the potential to reach SDGs 3, 11, and 13 when accompanied by policy, funding, and inclusive planning.

### 3. Equity, Inclusion, and Disability-Centred Design

The review also examined whether digital interventions appropriated the concerns of disabled and marginalised people. Multiple studies revealed that adequately designed digital tools had the potential to decrease disparities through the elimination of mobility, stigma, and isolation barriers. However, disability inclusion in design varied widely.

Ugwu et al. (2025) have presented a case of equity-based digital health in conflict-related LMICs, which prevent access gaps by utilizing community-grounded transportation models (23). The case presented by Khuld (2024) showed that the deployment of female providers, multilingual apps, and cross-cultural understanding by Sehat Kahani increased the number of underserved women, children, and elderly patients receiving care (26).

An example of a disability-focused case was that of Pasipanodya and Shem (2020), who used the case of a spinal cord injured individual who received access to rehabilitation during wildfires. This reiterated the need of anticipatory, inclusive planning (25). Conversely, Emuk et al. (2023) warned that the failure of infrastructure and a lack of digital literacy may increase the disparities among the vulnerable groups, including the ones, who may require frequent rehabilitation (22).

Bell et al. (2017) and Jones et al. (2020) examined treatment of mental health issues in veterans and military members both of whom experience issues with PTSD and depression. They determined that the digital supply lessened stigma and expanded access (24, 28). Yet, Jones et al. concluded that no study had examined the influence of race, gender, or culture on user experiences (24).

Generally, these results highlight the need to ensure that digital health is proactively inclusive and rights-based. In the absence of such design, it will be all too easy to extend inequities on the system to the digital realm through interventions.

### 4. Barriers and Enablers to Implementation

Common barriers and facilitators among all of the studies were identified, to inform digital health implementation barriers. Disaster areas were a significant problem to the infrastructure because of electricity and internet blackouts (22, 26). Patients and provider digital illiteracy added another constraint to the adoption of tools.

The issue of privacy and information security also came up. Khuld (2024) made sure that HIPAA is complied with, however, others (24, 28) also found it difficult to control user safety and confidentiality, particularly when they rush to provide remote trauma services.

Notable enablers were pre-existing digital systems, exemplified in the teleSCI programme. The funding, expansion, and organization occurred through cross-sector collaborations, i.e., between the government, NGOs, and tech companies (23, 26). Uptake and trust were also encouraged by technology localisation e.g. culturally sensitive platforms and female-led teams.

To conclude, innovation is not the only thing that can help digital health-related solutions to succeed in a crisis environment; it must be accompanied by inclusive planning, ethical protection, and institutional reinforcement. Strategic action against barriers is an essential element of the development of inclusive and resilient care systems.

## Discussion

### Reframing Digital Health in Crisis- Promise, Pitfalls and Patterns

The findings of this scoping review point to a growing global recognition of digital health as an adaptive mechanism for delivering essential care in contexts where traditional health systems are disrupted by natural disasters or conflict. With interventions ranging between mobile telemedicine in the case of floods to virtual therapy in the case of traumatized veterans, the interventions identified here show how digital solutions can address vulnerable populations in cases where physical infrastructure is compromised.

Nonetheless, the majority of reviewed interventions were reactive, responding in short-term rather than being part of the systemic resilience of health systems over an extended period. Considering the example of the Sehat Kahani model (Khuld, 2024), scalable care during floods was provided but it is unclear how this model can be sustained in the long run through ideally institutionalisation or otherwise adaptation in non-emergency periods (26). This tendency indicates the fact that digital health remains a subject to contingency despite the current shift towards normalization (29).

Although numerous included studies in this review, reported to target a group described as vulnerable or marginalised, a shallow treatment was evident of the reality of vulnerability in multiple layers, especially in the case of people with disabilities. In many cases, vulnerability was treated as a homogenous category, without accounting for how disability, gender, age, poverty, and geography intersect to shape access to care (30). This limits the extent to which these digital solutions can be described as inclusive or transformative (31).

Notably, as far as digital health interventions are concerned, their effectiveness is severely dependent on the local context especially infrastructure, governance, and digital literacy (32, 33). Even well-developed digital systems are liable to malfunction in delicate settings already aroused by conflict or climatically partnered disasters. The robustness of digital health tools cannot be decoupled with the robustness of the environments they rely on (34, 35).

### Equity Paradox and Digital Health

Even though there was a lot of intervention to allow previously excluded groups, including those with disabilities, women in rural settings, or displaced population groups, the actual scenario was not ideal. Although models such as Sehat Kahani showed gender-sensitive designs, such as women-run virtual clinics or multilingual dashboards, majority of the interventions did not specify how the services were modified to accommodate the varied needs of the disabled users (26).

This points to a broader issue: the lack of disaggregated data and participatory design. None of the studies included in this review reported outcome data separately for disabled users, and very few involved users from marginalised groups in the planning or evaluation of interventions. Without this level of detail, it becomes difficult to know whether digital tools are actually reaching those most at risk, or simply improving access for those who are already digitally connected.

Further, digital access per se (devices, internet, personal space) cannot be relied upon in disaster- or conflict zones. Some research accepted the existence of these barriers; however, not many provided tangible results. In a study by Emuk et al (2023), tele rehab was marred due to power failure and unstable internet after the earthquake. These are not marginal obstacles but they determine who is eligible and who is marginalized (22).

Equitable design will not be limited to presenting a digital solution, but will become inclusive design in every point including interface, staffing, training, and follow-up. Most of the reviewed interventions failed to take into account such practical considerations, leading to systems that subsequently alienate the same populations that they are supposed to serve (36).

### Causation and Causation: Structural Gaps and Sustainability

One of the main findings of the review is delivering no evidence on long-term sustainability and systemic integration. A majority of the interventions were performed as an emergency measure and evaluated on the short-term basis. These observations, though helpful, do not provide an in-depth clarity into the long-term performance of digital platforms and specifically in environments, such as the ones already experiencing under-resourcing of the system, or the political imbalance.

As an illustration, models proposed by Mesmar et al. (2016) and Lokmic-Tomkins et al. (2024) were implemented in unstable or weakly provided territories (21, 27). Although they temporarily alleviated service gaps, there was no indication of whether they led to permanent results in terms of workforce capacity, policy, or integration. Unless there is sustainable financing or national-scale convergence, there is a danger that they will simply end up as defunct pilot projects (37).

It is important to note that only a single study was high quality randomised controlled trial (28). This limits our understanding of what works, for whom, and in what contexts. The lack of consistent reporting on user satisfaction, clinical outcomes, or cost-effectiveness also makes it difficult to compare models or scale successful ones across regions (38). In other words, while the potential of digital health is well-documented, the evidence of its effectiveness in fragile settings remains uneven and difficult to generalise (34).

Another gap is the lack of digital health in comprehensives emergency preparation systems. Such researchers as Pasipanodya and Shem (2020) were among the few who noted the promise of prior infrastructure in case of sudden-onset disaster. The majority of others were the ad-hoc responses, which indicate that the digital health could not be integrated into national crisis plans (25).

Digital tools cannot just support resilience effectively, as envisioned in SDGs 3, 11 and 13, but must be part of their structure - policy, budgeting, training, and workflows in health systems.

### Policy, Practice, and Research Implications

The review has a number of important implications. Humanitarian actors and policymakers should not consider digital health a supportive intervention but an essential crisis care strategy. It is important to be integrated in the disaster preparedness plans and should be co-ordinated with the existing health systems (39).

Secondary, digital health initiatives are forced to develop beyond simply allowing access but to inclusion. Inclusion does not only mean availability but proactive accommodation to the requirements of users with disabilities, low literacy, or sub-optimal connectivity. It must have offline capability, audio-visual interfaces, and culturally specific information. Besides, the design and evaluation processes need to involve affected communities including persons with disabilities (40).

More specific evidences are also required, and they should be stronger. Not many studies applied rigorous or longitudinal methodologies, and data were rarely disaggregated by either gender, disability, or geography. Future studies in this area should adopt implementation and realist evaluation studies to aim at determining what, and in whom, and under what circumstances things do or do not work. Data privacy and informed consent are also ethical considerations, which should be central to data sharing, especially in place with low governance.

Though this review was concentrated on crisis situations, its implications are applicable to the global digital health policy. The pandemic of COVID-19 exhibited the potential, and the pitfalls of quick widespread digital health adoption. Policy should use lessons learned elsewhere in unsafe environments to make sure that innovation does not widen pre-existing disparities (41).

## Limitations

This review has several limitations that should be acknowledged. First, although the search was comprehensive and included multiple databases, relevant studies published in non-indexed sources or grey literature may have been missed. This is particularly relevant in humanitarian settings, where many impactful interventions are documented in project reports or unpublished evaluations rather than peer-reviewed journals.

Second, the review included studies with a variety of designs, ranging from narrative reviews to case studies and one clinical trial. While this diversity reflects the early and applied nature of the field, it also limits the ability to draw strong conclusions about effectiveness or causality. The absence of robust quantitative evaluations in most studies means that many of the findings remain descriptive and context-dependent.

Third, while the review aimed to assess digital health interventions for vulnerable and disabled populations, few studies disaggregated their findings to reflect these specific subgroups. This limited our ability to evaluate the impact of interventions on persons with disabilities in particular. Additionally, most studies focused on interventions during or after emergencies, with very little attention paid to preparedness or long-term integration into health systems..

## Recommendations

According to the results of this review, there are a number of areas that researchers, policymakers, and implementers should pay attention to. The constraints with respect to conducting more rigorous and context-sensitive studies and methodologies (mixed-methods and longitudinal study designs), which are beyond the descriptive reporting, are pressing in the future. There must be the standardization of disaggregated outcome data, especially in terms of disability, gender bias, age and socioeconomic status to make sure that digital interventions not only become widely dispersed but are equally efficient in their effects. Moreover, participatory methods should be the priority as they allow including people with disabilities and other neglected groups in the process of designing, implementing, and testing digital health technologies. This would assist in designing interventions which meet the practical requirements of the user in crisis contexts. They are also encouraging governments and humanitarian organisations to incorporate digital health as part of the disaster preparedness regime as opposed to reactionary deployments. The policies must coordinate funding, personnel development, moral protection, and interoperability guidelines throughout the systems to favour sustainability. And last, evidently there is a requirement of more investment in implementation research to look beyond simple question of whether digital health tools work but under what circumstances they succeed or fail. In the absence of such evidence, the potential role of digital health in enabling inclusive and resiliency in care system in disaster and conflict settings will continue to go under-realised.

## Conclusion

This scoping review outlines the changing scenario of digital health in assisting vulnerable and disabled groups in natural disasters and wars. Although the interventions considered have a distinct optimization potential in terms of access, continuity of care, and adaptability of the system, the evidence base is discontinuous and inconsistent. A significant majority of studies relied on short-term implementation and could not pay much attention to long-term sustainability, structural inclusion, or disability-related outcomes.

Notwithstanding these shortcomings, the review indicates that, when inclusive and built with the possibilities of integrating them into the intended system, digital health interventions can make a positive contribution to the objectives of SDGs 3, 10, 11, and 13. Nevertheless, this will necessitate a conscious movement towards inclusive and equity-based strategies enshrined into national preparedness plans rather than adopting reactive measures. The argument in support of robust and accessible digital health systems has never been stronger in times of increased and compounded global crises.

## Data Availability

All data underlying the results are presented in the manuscript and its Supporting Information files, and no additional source data are required.

## Declarations

### 1: Ethics approval and consent to participate

Not applicable. No human participants, human data, and human tissue were directly collected by the authors in this scoping review.

### 2: Publication consent

Not applicable. No data of any single individual was included in this manuscript.

### 3: Data and materials availability

This published article includes the data generated and analysed during this scoping review. One search strategy and retrieved study characteristics have been provided as tables in the manuscript. The supplementary file contains screenshots from all the databases used for this scoping review.

## References

1. Ko J, Lee HF, Leung CK. War and warming: The effects of climate change on military conflicts in developing countries (1995–2020). Innovation and Green Development. 2024;3(4):100175.

2. Abbass K, Qasim MZ, Song H, Murshed M, Mahmood H, Younis I. A review of the global climate change impacts, adaptation, and sustainable mitigation measures. Environmental Science and Pollution Research. 2022;29(28):42539–59.

3. Herre B, Rodés-Guirao L, Roser M. War and peace. Our World in Data. 2024.

4. Vuong Q-H, Nguyen M-H, La V-P. The overlooked contributors to climate and biodiversity crises: military operations and wars. Environmental Management. 2024;73(6):1089–93.

5. Kutcher S, Borisch B. Health and global security: challenges and considerations. Springer; 2025.

6. Jha MK, Dev M. Impacts of climate change. Smart internet of things for environment and healthcare: Springer; 2024. p. 139–59.

7. Martins FP, Paschoalotto MAC, Closs J, Bukowski M, Veras MM. The double burden: climate change challenges for health systems. Environmental health insights. 2024;18:11786302241298789.

8. Paz S, Díaz J, Negev M, Linares C. Climate change and global health. Handbook of Epidemiology: Springer; 2024. p. 1–35.

9. Mosleh M, Al Jeesh Y, Dalal K, Eriksson C, Carlerby H, Viitasara E. Barriers to managing and delivery of care to war-injured survivors or patients with non-communicable disease: a qualitative study of Palestinian patients’ and policy-makers’ perspectives. BMC Health Serv Res. 2020;20(1):406.

10. Tanggono A, Prajnaparamita I, Nastaghfiruka P. The Role of Technology in Enhancing Public Health Preparedness for Disasters. Riwayat: Educational Journal of History and Humanities. 2025;8:1134–46.

11. Stoumpos AI, Kitsios F, Talias MA. Digital Transformation in Healthcare: Technology Acceptance and Its Applications. Int J Environ Res Public Health. 2023;20(4).

12. Friedman C, VanPuymbrouck L. Telehealth Use By Persons with Disabilities During the COVID-19 Pandemic. Int J Telerehabil. 2021;13(2):e6402.

13. Al Shami A, Nashwan AJ. Introduction to Disability in Low-and Middle-Income Countries. Global Health and Disability: Challenges in Low-and Middle-Income Countries: Springer; 2025. p. 1–11.

14. Farid ZI, Islam MA, Roberts PS, Glick J. Disability Inclusive Disaster Risk Reduction (DIDDR) in South Asia: Status, Prospects, and Challenges. 2024.

15. Reduction UNOfDR. 2023 Global Survey Report on Persons with Disabilities and Disasters. 2023.

16. Cooper A. Vulnerable populations in disasters: Health effects and needs. 2017.

17. Sylla B, Ismaila O, Diallo G. 25 Years of Digital Health Toward Universal Health Coverage in Low- and Middle-Income Countries: Rapid Systematic Review. J Med Internet Res. 2025;27:e59042.

18. Zia A. Digital Health for Vulnerable and Disabled Populations in Natural Disaster and Conflict Settings: A Scoping Review Contributing to Sustainable Development Goals 3, 10, 11, and 13: OSF (Open Science Framework); 2025 [Available from: https://osf.io/y2ta3.

19. Levac D, Colquhoun H, O’Brien KK. Scoping studies: advancing the methodology. Implement Sci. 2010;5:69.

20. Tricco AC, Lillie E, Zarin W, O’Brien KK, Colquhoun H, Levac D, et al. PRISMA Extension for Scoping Reviews (PRISMA-ScR): Checklist and Explanation. Ann Intern Med. 2018;169(7):467–73.

21. Mesmar S, Talhouk R, Akik C, Olivier P, Elhajj IH, Elbassuoni S, et al. The impact of digital technology on health of populations affected by humanitarian crises: Recent innovations and current gaps. J Public Health Policy. 2016;37(Suppl 2):167–200.

22. Emük Y, Abasıyanık Z, Kahraman T. Telerehabilitation: A Promising Solution for Post-Earthquake Rehabilitation. İzmir Katip Çelebi Üniversitesi Sağlık Bilimleri Fakültesi Dergisi. 2023;8(2):647–51.

23. Ugwu CN, Ugwu OP, Alum EU, Eze VHU, Basajja M, Ugwu JN, et al. Sustainable development goals (SDGs) and resilient healthcare systems: Addressing medicine and public health challenges in conflict zones. Medicine (Baltimore). 2025;104(7):e41535.

24. Jones C, Miguel-Cruz A, Smith-MacDonald L, Cruikshank E, Baghoori D, Kaur Chohan A, et al. Virtual Trauma-Focused Therapy for Military Members, Veterans, and Public Safety Personnel With Posttraumatic Stress Injury: Systematic Scoping Review. JMIR Mhealth Uhealth. 2020;8(9):e22079.

25. Pasipanodya EC, Shem K. Provision of care through telemedicine during a natural disaster: a case study. Spinal Cord Ser Cases. 2020;6(1):60.

26. Khuld H. Impact of telemedicine mobile units on health outcomes, healthcare system resilience, and disaster response in flood-affected areas: a case study of Sehat Kahani. Interdisciplinary Journal of Epidemiology and Public Health. 2024;7(1).

27. Lokmic-Tomkins Z, Bhandari D, Bain C, Borda A, Kariotis TC, Reser D. Lessons Learned from Natural Disasters around Digital Health Technologies and Delivering Quality Healthcare. Int J Environ Res Public Health. 2023;20(5).

28. Bell KR, Fann JR, Brockway JA, Cole WR, Bush NE, Dikmen S, et al. Telephone Problem Solving for Service Members with Mild Traumatic Brain Injury: A Randomized, Clinical Trial. J Neurotrauma. 2017;34(2):313–21.

29. Dasgupta A. Adoption of digital health with a concept of patient as an organization 2022.

30. Kuran CHA, Morsut C, Kruke BI, Krüger M, Segnestam L, Orru K, et al. Vulnerability and vulnerable groups from an intersectionality perspective. International Journal of Disaster Risk Reduction. 2020;50:101826.

31. Xie Y, Fadahunsi KP, Kelleher C, Tarn DM, Grace A, J OD. Towards an inclusive digital health ecosystem. Bull World Health Organ. 2025;103(2):170–3.

32. Erku D, Khatri R, Endalamaw A, Wolka E, Nigatu F, Zewdie A, et al. Digital Health Interventions to Improve Access to and Quality of Primary Health Care Services: A Scoping Review. Int J Environ Res Public Health. 2023;20(19).

33. Fitzpatrick PJ. Improving health literacy using the power of digital communications to achieve better health outcomes for patients and practitioners. Front Digit Health. 2023;5:1264780.

34. Mumtaz H, Riaz MH, Wajid H, Saqib M, Zeeshan MH, Khan SE, et al. Current challenges and potential solutions to the use of digital health technologies in evidence generation: a narrative review. Front Digit Health. 2023;5:1203945.

35. Ahmed SK, Hussein S, Chandran D, Islam MR, Dhama K. The role of digital health in revolutionizing healthcare delivery and improving health outcomes in conflict zones. Digit Health. 2023;9:20552076231218158.

36. Bucher A, Chaudhry BM, Davis JW, Lawrence K, Panza E, Baqer M, et al. How to design equitable digital health tools: A narrative review of design tactics, case studies, and opportunities. PLOS Digit Health. 2024;3(8):e0000591.

37. Bertone MP, Jowett M, Dale E, Witter S. Health financing in fragile and conflict-affected settings: What do we know, seven years on? Social Science & Medicine. 2019;232:209–19.

38. McKeever L. Overview of Study Designs: A Deep Dive Into Research Quality Assessment. Nutr Clin Pract. 2021;36(3):569–85.

39. Doarn C, Merrell R. Telemedicine and e-Health in Disaster Response. Telemedicine journal and e-health : the official journal of the American Telemedicine Association. 2014;20.

40. Townley C, Koop C. Exploring the potential and limits of digital tools for inclusive regulatory engagement with citizens. Government Information Quarterly. 2024;41(1):101901.

41. Getachew E, Adebeta T, Muzazu SGY, Charlie L, Said B, Tesfahunei HA, et al. Digital health in the era of COVID-19: Reshaping the next generation of healthcare. Front Public Health. 2023;11:942703.

